# Seasonal variation in SARS-CoV-2 transmission in temperate climates

**DOI:** 10.1101/2021.06.10.21258647

**Authors:** Tomáš Gavenčiak, Joshua Teperowski Monrad, Gavin Leech, Mrinank Sharma, Sören Mindermann, Jan Markus Brauner, Samir Bhatt, Jan Kulveit

**Affiliations:** Epidemic Forecasting; Future of Humanity Institute, University of Oxford; Faculty of Public Health and Policy, London School of Hygiene and Tropical Medicine, UK; Department of Health Policy, London School of Economics and Political Science, UK; Department of Computer Science, University of Bristol, UK; Department of Statistics, University of Oxford, UK; Department of Engineering Science, University of Oxford, UK; Oxford Applied and Theoretical Machine Learning (OATML) Group, Department of Computer Science, University of Oxford, UK; Department of Public Health, University of Copenhagen, Denmark

## Abstract

While seasonal variation has a known influence on the transmission of several respiratory viral infections, its role in SARS-CoV-2 transmission remains unclear. As previous analyses have not accounted for the implementation of non-pharmaceutical interventions (NPIs) in the first year of the pandemic, they may yield biased estimates of seasonal effects. Building on two state-of-the-art observational models and datasets, we adapt a fully Bayesian method for estimating the association between seasonality and transmission in 143 temperate European regions. We find strong seasonal patterns, consistent with a reduction in the time-variable *R*_*t*_ of 42.1% (95% CI: 24.7% – 53.4%) from the peak of winter to the peak of summer. These results imply that the seasonality of SARS-CoV-2 transmission is comparable in magnitude to the most effective individual NPIs but less than the combined effect of multiple interventions.

## 1. Introduction

Since the onset of the COVID-19 pandemic, the role of seasonal variation in SARS-CoV-2 transmission has received significant scientific and political attention [1]. Understanding seasonal patterns is vital, as it enables more accurate inferences about current trends in transmission and how they may change over the longer term. For example, a proper understanding of seasonality can help policymakers avoid attributing declining incidence over the summer to population immunity alone, when in fact seasonality may be playing a meaningful role.

While seasonal variation is well-established for many respiratory viral infections [2], and some studies have suggested associations between temperature, humidity, and COVID-19 incidence [3, 4, 5, 6, 7], other analyses have failed to show a robust role of climate and weather [8, 9], particularly when population immunity is low [10]. A recent review found that the evidence remains inconclusive [11].

A further complication is that temperature, humidity, and UV radiation plausibly affect transmission and incidence through a range of biological and epidemiological mechanisms [2, 12]. These include virus stability and viability [13, 14], host susceptibility and immune response [15, 16], human behaviour [17, 18], and social factors such as holidays and school calendars [19, 20]. This multitude of plausible causal pathways makes it exceedingly difficult to disentangle the influence of various seasonal factors, particularly given the extensive multi-collinearities and interactions between environmental, biological, and behavioural elements [21, 22]. Appendix A shows an overview of the various causal pathways, including existing literature and evidence on the collinearities between various factors. As Lofgren and colleagues note in the context of influenza, “the myriad theories accounting for seasonality (…) suggest that the elegant and predictable periodicity of nonpandemic influenza is caused by a less-than-straightforward interaction of many different factors,” meaning that “recognition of this complexity, as well as the likelihood that seasonality arises from many different factors, is essential for continued examination and elucidation of seasonality” [17].

Given the severe methodological challenge of disentangling these interrelated factors, a more tractable solution is to approach seasonality holistically with the purpose of understanding its overall effects. In this study, we infer a single seasonality parameter, describing the amplitude of the yearly variation in the time-variable reproduction number, *R*_*t*_, for one climate region. While this single parameter does not disentangle the individual effects that comprise a seasonal profile, it accounts for the overall magnitude of the seasonal effect on SARS-CoV-2 transmission and thereby provides valuable insights for long-term policy planning.

Since both COVID-19 incidence and the presence of governmental non-pharmaceutical interventions (NPIs) have waxed and waned in consecutive waves since early 2020, adjusting for NPI effects is crucial for any effort to infer the influence of seasonality on transmission, yet early analyses of environmental drivers have largely not done so [11, 7, 5].

Some studies of environmental factors have taken indirect approaches to avoiding the influence of NPIs on their environmental estimates. In a recent study of the association between humidity, temperature, and SARS-CoV-2 transmission in Europe and North America, Landier and colleagues exclude from their analysis any periods at least 28 days after the implementation of “lockdown measures” [4]. Ma and colleagues do include periods where measures are in place, however, instead of directly utilising data on NPIs, they use smoothed spatial and temporal splines to indirectly adjust for their influence [3]. Smith and colleagues compare the role of temperature in the presence and absence of “lockdown” in the United States but only include a binary measure of whether stay-at-home orders were in place [22].

By contrast, we directly adjust for the influence of specific interventions by extending two hierarchical Bayesian models of NPI effects from Brauner et al. [23] and Sharma et al. [24] to include a term representing the multiplicative seasonal influence on the effective reproduction number.

Employing a common technique in infectious disease modelling [25, 26], we assume the seasonal variation itself is described by a sinusoidal modulation. We re-analyse data from existing studies [23, 24] while restricting the scope to European regions in the temperate climate zone, where we assume the seasonality effect to be comparable both in its environmental and behavioural causal components.

## 2. Methods

Published hierarchical Bayesian models of NPI effects [23, 24, 27] typically assume the time-variable reproduction number *R*_*t*_ to be a product of *R*_0_, the “natural” reproduction rate without mitigations, multiple terms representing the effects of interventions, and noise terms modelling growth speed variance, of the form:

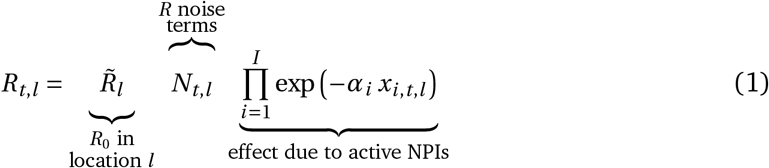

where exp(*−α*_*i*_) is the estimated effect of intervention *i* and *x*_*i,t,l*_ are indicator variables of interventions implemented on a given date in a given location. Daily reproduction rates are then typically connected to observed data on cases or deaths by growth rates, compartmental epidemiological models, renewal processes, etc. The noise term *N*_*t,l*_ then varies with the model, e.g. a log-normal multiplicative factor in Brauner et al. [23] and a random walk-based multiplicative factor in Sharma et al. [24]. Mechanistically, the noise term can be intuitively thought of as a random effect that accounts for residual variation not captured by the NPI effects.

To account for seasonality, we substitute *R*_*t,l*_ with 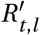 (adjusted for seasonality) and let the model infer a single seasonality amplitude parameter *γ* along with its other parameters. This minimal modification aims to preserve the demonstrated robustness of the original models [23, 24, 28]. The final seasonality amplitude estimate is then pooled from the two models, equally sampling from their posterior distributions. Note the seasonal adjustment to *R*_*t*_ is *shared* across all locations *l* and therefore captures common dynamics between locations not explained by the location specific noise terms or NPIs.

We model seasonality as a sinusoidal multiplicative factor Γ(*t*) to *R*:

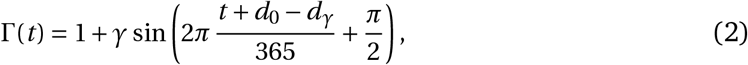

where *γ* is the intensity (amplitude) of the seasonal effect, *d*_*γ*_ is the day of the year of the highest seasonal effect on *R*, and *d*_0_ is the first day of the respective dataset. We assume a single, common seasonal effect for countries in similar climates and relative proximity along dimensions such as income, political structure, and culture. While average temperatures clearly are different between countries within the region, with a strong dependence on latitude, the amplitude of the seasonal variation is assumed to be similar. For both models, the time and location-specific *R*_*t,l*_ is replaced with seasonal 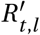:

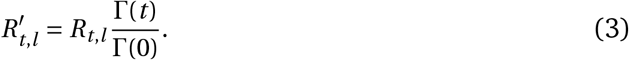

Note that we divide Γ(*t*) by Γ(0) to have Γ(*t*)/Γ(0) *=* 1 at *t =* 0 and 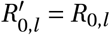, i.e. the seasonality multiplier is normalised to 1 at the start of the window of analysis. This means that the priors over 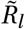 need not be adjusted in either model. For both models, we assume an uniform prior *γ ∼U* (*−*0.95, 0.95)^1^.

The amplitude of the cyclical seasonal variation (*γ*) can be converted to the reduction in transmission associated with going from the peak of winter to the peak of summer (i.e., peak-to-trough) as 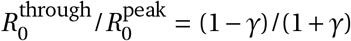. Similarly, the amplitude can be directly converted to the expected reduction between adjacent seasons such as peak winter to mid-spring or mid-spring to peak summer (i.e., peak-to-mid).

Our analysis utilises January 1 as the seasonal peak day *d*_*γ*_, as this date is both close to the center of a stable range of *d*_*γ*_ in the sensitivity analysis of Appendix F.2, as well as close to January 3, the median peak date inferred by a model with variable *d*_*γ*_ in Appendix F.1. Note that while we show January 1 to be a robust choice of *d*_*γ*_, we are not aiming at determining its exact value.

Our analysis relies on the original data from Brauner et al. and Sharma et al. [23, 24]. We restrict the dataset of Brauner et al. to the 29 countries in the temperate Europe region (see Appendix B) and use the Sharma et al. dataset without any modifications, for a total of 143 regions of analysis. We follow the same pre-processing steps as in the original datasets, in particular, we exclude any data points where the prevalence of variants of concern exceeded 10% during a given day, to mitigate potential bias introduced by more transmissible strains.

## 3. Results

Using two model structures and datasets on non-pharmaceutical interventions covering 72% of the 2020-2021 period in Europe, we estimate the seasonality parameter *γ* and the time-variable seasonal multiplier Γ(*t*) (Figure 1).

**Figure 1:**
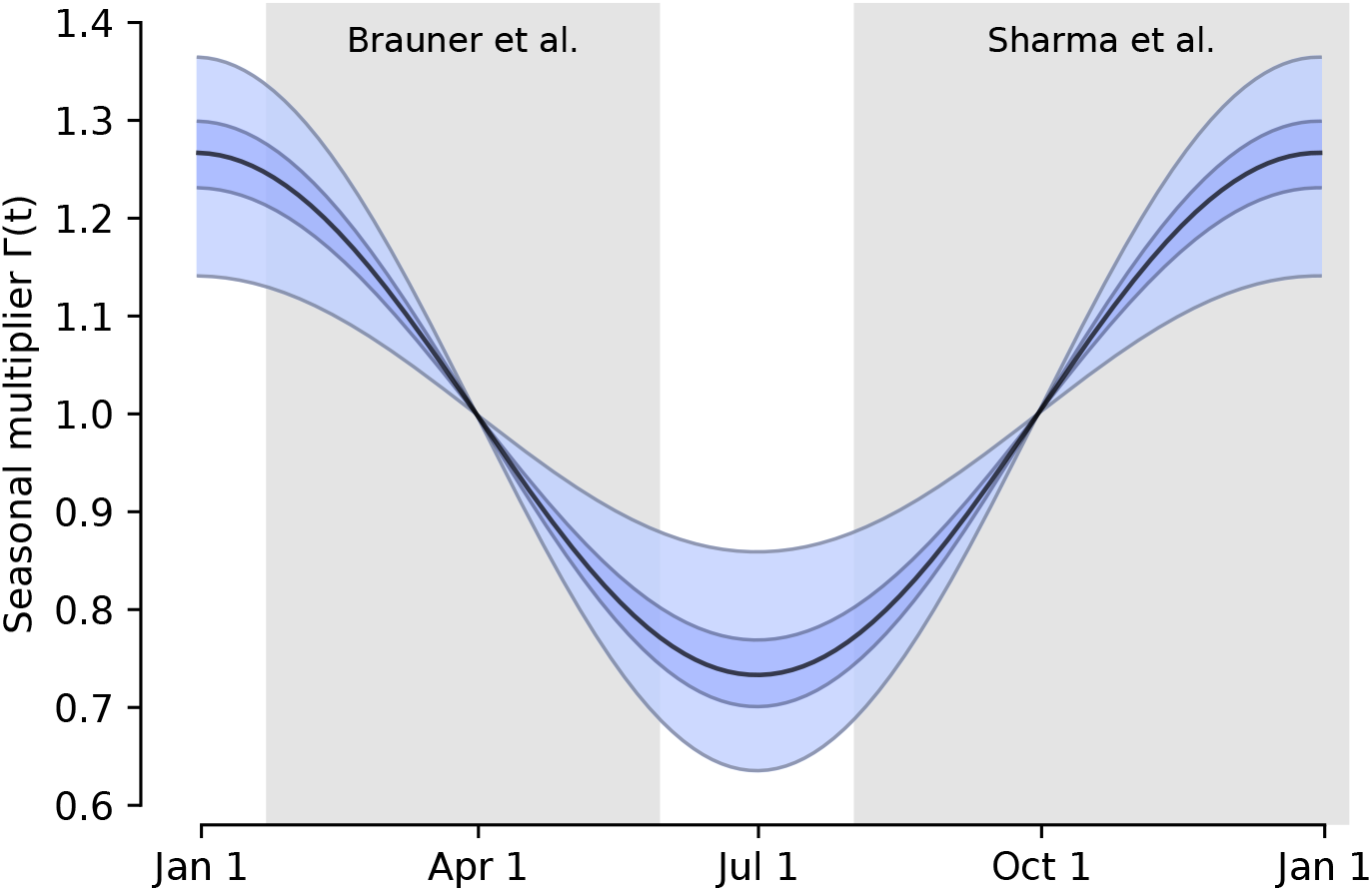
The inferred seasonal *R* multiplier Γ(*t*) of the combined models estimate, with 50% and 95% confidence intervals. Gray boxes indicate data range of each dataset.

Our combined estimates from the two models are consistent with a reduction in *R* of 24.7% to 53.4% (95% CI) from January 1, the peak of winter to July 1, the peak of summer, with a median reduction of 42.1% (Figure 2 and detailed results in Appendix C).

**Figure 2:**
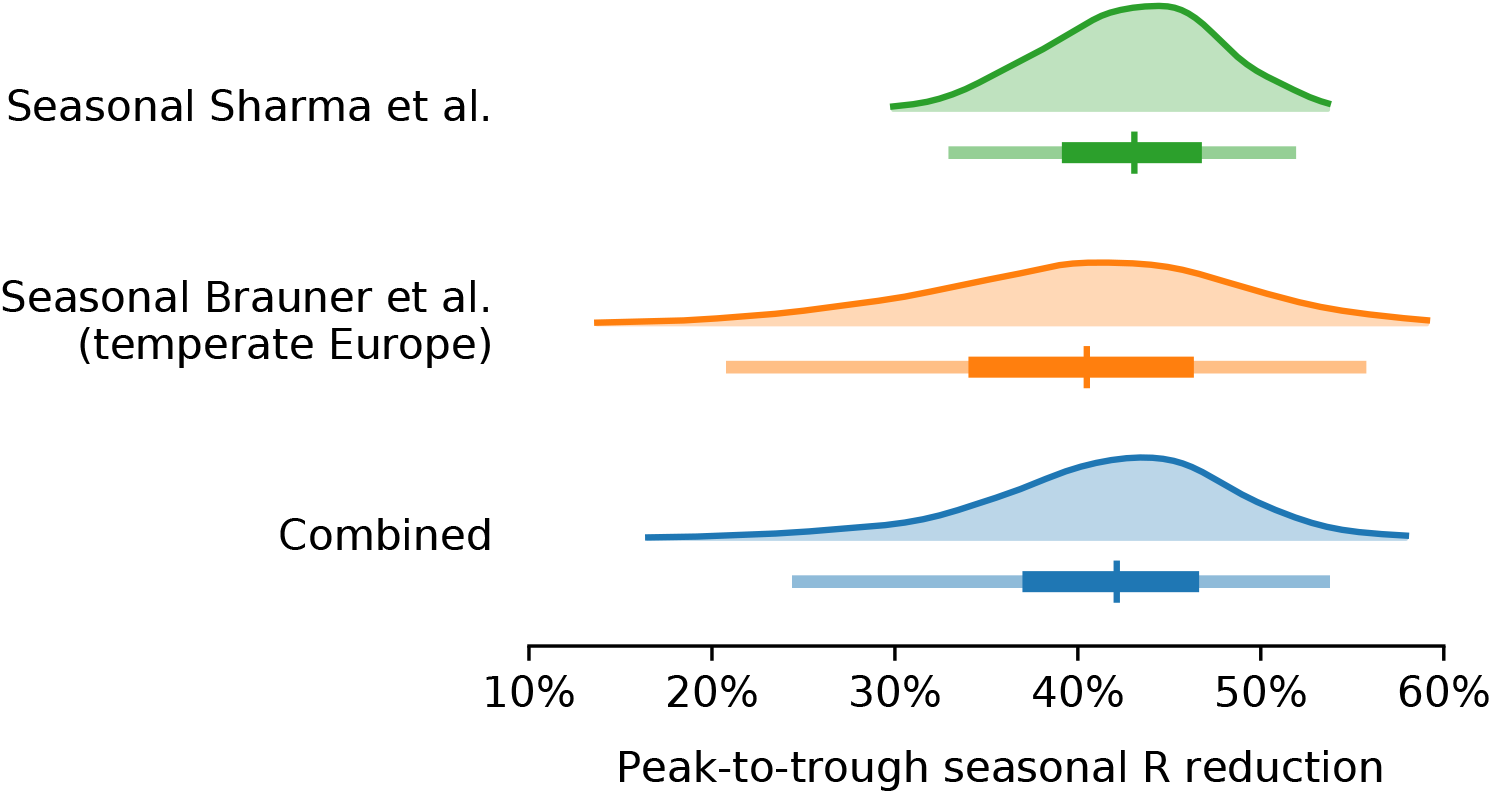
Posterior distributions of the *R* reduction on July 1 relative to January 1 with median, 50% and 95% confidence intervals.

Modelling seasonality alongside non-pharmaceutical interventions allows us to gain a sense of the epidemiological importance of environmental factors. We find that the transition from winter to summer is associated with a reduction in transmission that is comparable to or greater than the effects of individual interventions, but less than the total effect of combined interventions (Figure 3).

**Figure 3:**
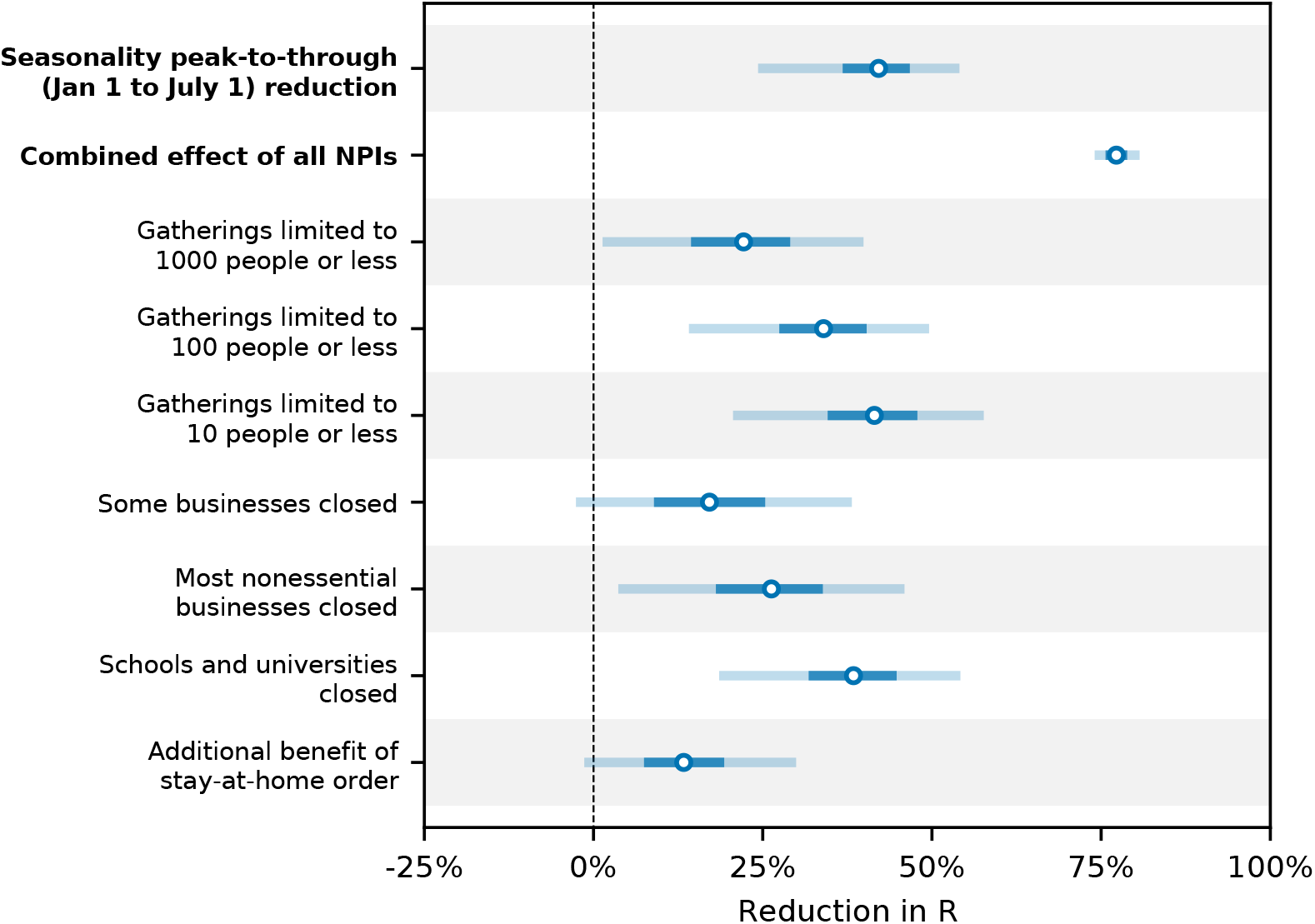
Comparison of the inferred peak-to-trough *R* reduction effect of seasonality (combined from both models) to the NPI reductions inferred by Brauner et al. [23], with 50% and 95% CIs. The seasonal effect is lower than the combined NPI effect but higher than or comparable to the individual NPI effects.

Figure 3 compares our seasonality estimates to the original effect estimates from Brauner et al., as their robustness is well-established [28, 23]. Although these estimates are based on analysis that included countries outside temperate Europe, we find that restricting our analysis to temperate regions has little effect on the inferred total effect of NPIs and thus should not invalidate the comparison (Appendix F.4).

Beyond NPIs, voluntary changes in behaviour and contact patterns constitute important influences on the reproduction rate. As noted, seasonal variation in behavioual patterns such as time spent indoors are an important component of our holistic conception of seasonality (Appendix A). However, if there are behavioural changes over time that are causally unrelated with the transition between seasons, these may be mistakenly attributed to a causal effect of seasonal forcing. If, for example, voluntary personal protective behaviours beyond compliance with NPIs increased during the spring and decreased during the autumn, this would provide an alternative explanation of the respective fall and rise in transmission during those seasons. To examine whether behavioural changes over the course of the pandemic influenced our results, we incorporated mobility in the model by treating country-level reductions in mobility as a distinct NPI (see Appendix D). We find that our seasonality estimates are robust to adjusting for mobility trends. Specifically, our estimate of *γ* changes by less than 1.64% with the adjustment for mobility. While this finding strongly suggests that the observed seasonal pattern cannot be explained by unrelated changes in behaviour, it should be noted that data on mobility may not capture every relevant aspect of behavioural trends and that we therefore cannot conclusively rule out the possibility that our estimates are influenced by changes in behaviour.

Incorporating seasonality into models of NPI effectiveness may also improve their estimates by explaining residual variation in the inferred reproduction rate. A key advance in the model proposed by Sharma et al. was the incorporation of a stochastic random walk process on the basic reproduction number to flexibly account for trends in transmission due to unobserved factors [24]. We find that including the seasonality term reduces the magnitude and asymmetry of the random walk considerably, thereby reducing the internal model variation (Appendix E). Specifically, we find that the mean square displacement (MSD) of the random walk in log-space is 0.131 for the non-seasonal model and 0.072 for the seasonal model. These results suggest a considerable amount of the residual variation can be explained by a common seasonality profile.

Estimates of seasonality and NPI effects are sensitive to modelling choices [23, 28, 24]. It is therefore vital to include a sensitivity analyses of free parameters and inputs to ensure consistent results. Relying on the demonstrated robustness of the original models, we focus primarily on the parameter that we introduce in the form of peak seasonality day. We find that the inferred mean peak-to-trough reduction in *R* varies by less than 5% across all the analysed peak seasonality dates in December and January (Appendix F.2). Although the seasonality magnitude is somewhat sensitive to setting the winter peak to different dates in February, these dates are considerably later in the year than the median peak date inferred in our sensitivity analysis, January 3 (see Appendix F.1).

Since the seasonality term we introduce is directly related to *R*_*t,l*_ through Equation (3), we also examine the sensitivity of our results the mean initial *R*_0_ prior. We find that our results are robust to univariate variation in this parameter, with the seasonal Sharma et al. model being the most sensitive (Appendix F.3).

## 4. Discussion

The clear seasonal patterns of other respiratory viruses give us strong prior reasons to expect seasonal variation in SARS-CoV-2 transmission [2], and the strong associations we observe in temperate Europe match this expectation. While reductions in reproduction rates and case numbers are not directly comparable, another recent analysis by Chen et al. [7] infers a 64% reduction in cases from one season to the next based on a cross-sectional regression at a single point in time, similarly suggesting a significant role of environmental factors. The general magnitude of our results is also in line with previous assumptions about the magnitude of SARS-CoV-2 seasonal forcing. For example, Kissler et al. assume that the reduction of SARS-CoV-2 *R*_0_ between winter and summer peaks ranges from 10% to 40% [1], while Neher et al. assume values of *γ* between 0.3 and 0.7. Moreover, recent analyses have suggested a role of environmental factors in the B.1.1.7 lineage transmission intensity and that such factors may differentially affect the transmission of different variants of concern [6].

It is important to note that our results are not inconsistent with widespread outbreaks in warmer regions, nor do they imply that temperate regions cannot face surges in transmissions during summer periods. Despite moderate seasonal forcing, the time-variable reproduction number can remain well above 1 during the peak of the summer, particularly given high incidence of more transmissible variants such as lineage B.1.617.2 [29]. Indeed, in certain parts of Europe, *R* remained above 1 even during the warmer periods of the study window and transmission intensity currently remains high in several warmer regions across the world. Previous modelling has suggested that population immunity limits the role of environmental factors [10]. Consequently, vaccination rates, non-pharmaceutical interventions, and the prevalence of more transmissible variants will continue to be important determinants of transmission throughout the year.

Moreover, this study utilised variation in environmental and behavioural factors across time while holding the climate zone constant, and the observed results may not directly translate to comparisons across regions holding the season constant. In other words, the relationship between cooler periods and transmission within the temperate zone does not necessarily imply an exactly similar association between regional climate and transmission rates at any given point in time. This is because latitude is correlated with a wide range of epidemiological, demographic, and societal factors, each of which may affect transmission.

One major limitation of our analysis is that it relies on data from only one period of seasonality. We present the inferred seasonality estimates as the best estimate given the available data. Moreover, since our analysis focused exclusively on European regions in the temperate climate zone, the findings may not generalise to other climates, particularly as we have not identified the relative contributions of different causal mechanisms. Other respiratory infections show less seasonality in tropical regions relative to temperate regions as well as seasonal patterns with different peak timings, for example, during the monsoon season [2, 30]. Further research can shed light on the extent to which this is the case for SARS-CoV-2, and on the interaction between seasonality and latitude within climate regions.

More generally, this observational study demands caution when drawing conclusions about causality. Our analysis did not attempt to disentangle the various plausible causal pathways through which seasonality may affect transmission, and both environmental and behavioural factors can vary over the years. For example, behavioural patterns throughout the first year of the pandemic were likely exceptional, and while some behavioural changes are closely tied to modelled NPIs and thus do not bias our analysis, other relevant behavioural aspects may differ in subsequent years. Consequently, a granular focus on specific factors such as temperature, humidity, and behaviour is required for short-term prediction to inform policy.

Notwithstanding these limitations, the parsimonious seasonality form may be adequate to understand variations over time and aid long-term policy planning. Even without disentangling the underlying factors, incorporating seasonality can augment modelling efforts to more reliable anticipate changes in transmission patterns, particularly when adjusting for important factors such as non-pharmaceutical interventions.

For such forward-looking analyses of SARS-CoV-2 seasonality, it should be noted that our inferred seasonal associations do not include two factors that play significant roles in the seasonality of other respiratory viruses. First, we treat school closures, including for holidays, as NPIs in our model due to the role of closing educational institutions in the epidemic responses of many countries. This means that any effects of closing schools are attributed to the school NPI, rather than to seasonality. This is noteworthy considering that school calendars are considered an important driver of seasonality for other respiratory viruses [19, 31]. Consequently, the full extent of seasonality would likely be greater if it is construed to include school calendars. Second, the seasonal variation of some respiratory viruses, such as influenza, owes to a combination of both the direct seasonal forcing from biological and behavioural factors as well as the indirect influence of waning population immunity [32]. Given what is known about the robustness of acquired immunity within the first year of SARS-CoV-2 infection [33], the patterns we observe likely owe almost entirely to seasonal forcing. Going forward, the long-term seasonality of SARS-CoV-2 will depend in part on developments in population immunity as well as on the emergence of variants.

## 5. Conclusion

Failing to account for seasonality may lead to grave policy errors or a Panglossian outlook. For instance, a reduction in transmission over the summer may be misinterpreted as the result of herd immunity [34], and so lead to inadequate preparation for a resurgence during the colder months. Overestimating the role of environmental factors may be equally perilous, if policymakers anticipate a greater reduction due to seasonality than will actually occur.

## Data Availability

Spring 2020 dataset has been published in Brauner et al.: "The effectiveness of eight nonpharmaceutical interventions against COVID-19 in 41 countries".
Fall-winter 2020 dataset has been published in Sharma et al.: "Understanding the effectiveness of government interventions in Europes second wave of COVID-19".
The full implementation and all the datasets are publicly available on GitHub (links below).

https://github.com/gavento/covid_seasonal_Brauner

https://github.com/gavento/covid_seasonal_Sharma

## Acknowledgements

We thank Sebastian Funk, Adam Kucharski, and Swapnil Mishra for their especially insightful comments on the manuscript. We acknowledge the Department of Applied Mathematics of the Charles University in Prague for donating computational resources.

### Appendix A. Causal pathways for SARS-CoV-2 seasonality

#### Appendix A.1. Diagram of potential causal pathways

A complex web of environmental, biological, and behavioural factors contribute to the seasonality of respiratory viruses. Recent reviews by Moriyama et al. [2] and Tamerius et al. [35] provide discussions of the role that each of these factors play, with a particular focus on influenza. Figure A.4 illustrates some of the factors and potential causal pathways.

**Figure A.4:**
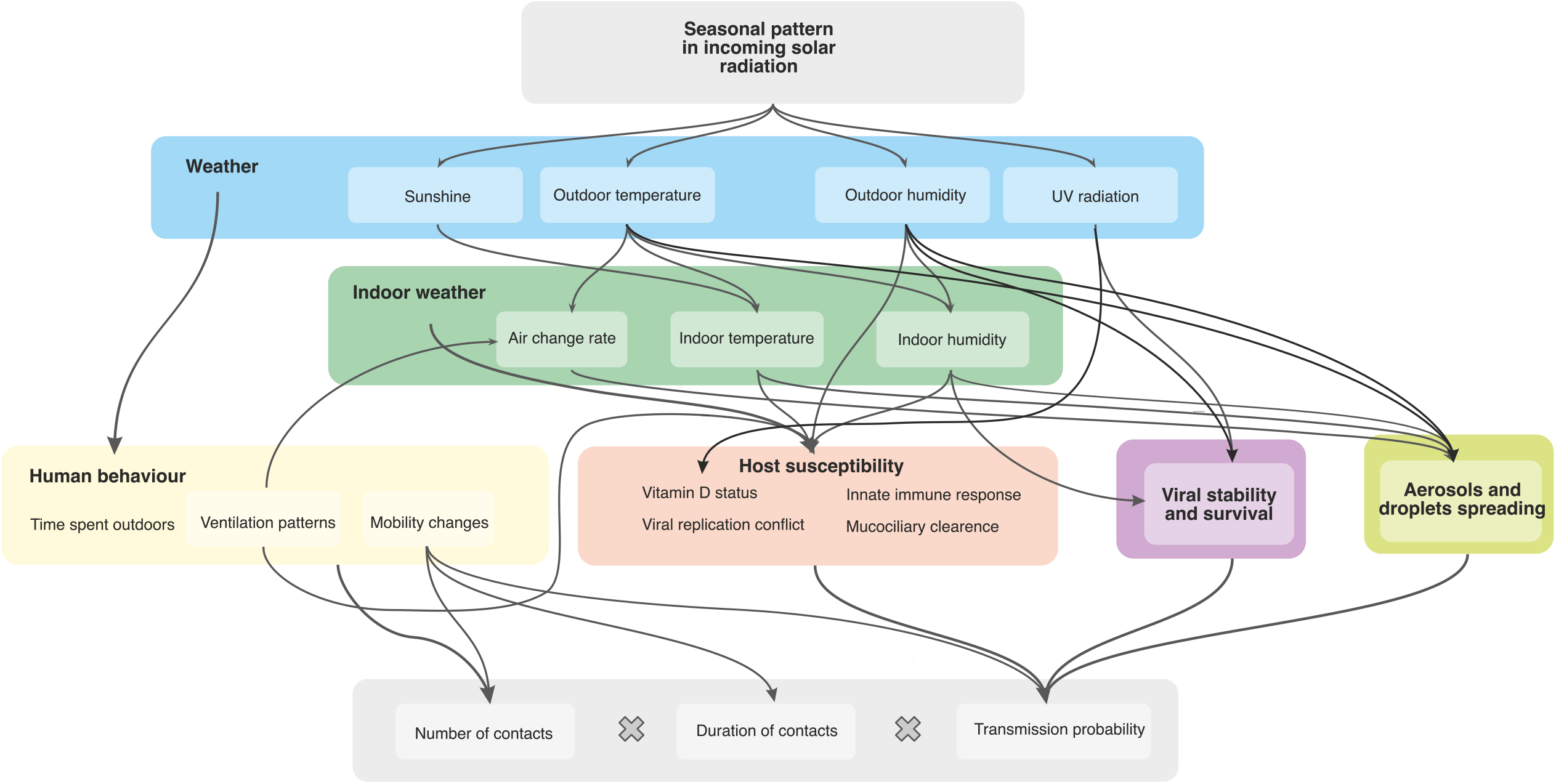
Diagram of proposed and established factors underlying seasonal patterns in viral transmission. Note that this diagram excludes school calendars, as these are subsumed under non-pharmaceutical interventions for the purposes of our analysis.

#### Appendix A.2. Existing evidence on causal pathways

For each of these pathways, theory and evidence have been presented in support of a causal relationship. However, extensive multi-collinearities and interactions complicate any effort to tease apart the exact contributions of different factors, particularly when considering population-level transmission dynamics where experimental approaches are intractable.

As Lipsitch and Viboud succinctly put it: *“Unfortunately, this potpourri of possible mechanisms places us in a kind of Popperian purgatory, in which data in support of every hypothesis exist, yet none of the hypotheses has been subjected to tests that are rigorous enough to reject it”* [21].

Table A.1 presents a (non-comprehensive) selection of evidence relating to some of the important causal pathways for viral seasonality.

**Table A.1:**
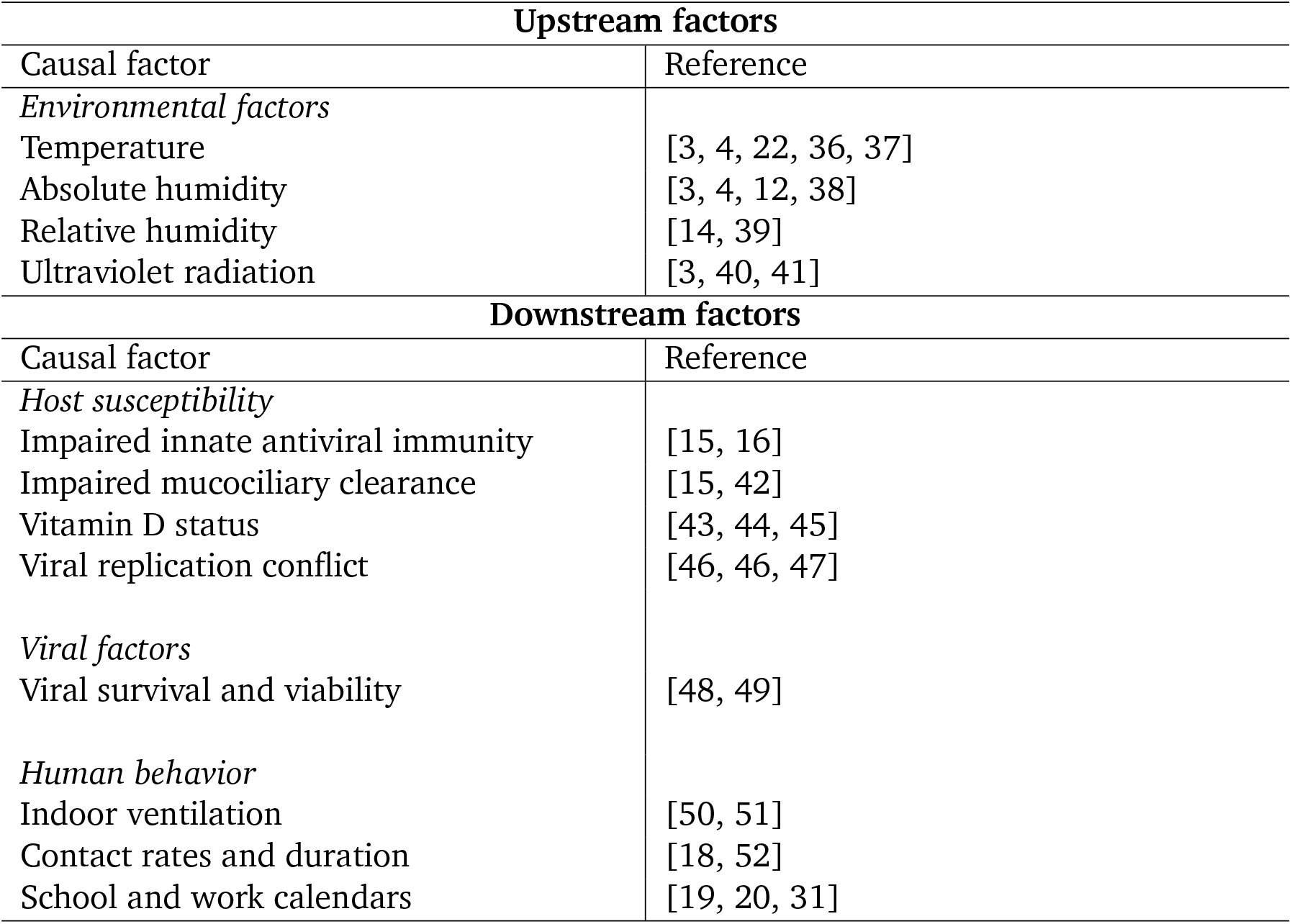
Selected evidence on factors driving respiratory virus seasonality.

| Upstream factors |  |
| --- | --- |
| Causal factor | Reference |
| *Environmental factors* |  |
| Temperature | [3, 4, 22, 36, 37] |
| Absolute humidity | [3, 4, 12, 38] |
| Relative humidity | [14, 39] |
| Ultraviolet radiation | [3, 40, 41] |
| Downstream factors |  |
| Causal factor | Reference |
| *Host susceptibility* |  |
| Impaired innate antiviral immunity | [15, 16] |
| Impaired mucociliary clearance | [15, 42] |
| Vitamin D status | [43, 44, 45] |
| Viral replication conflict | [46, 46, 47] |
| *Viral factors* |  |
| Viral survival and viability | [48, 49] |
| *Human behavior* |  |
| Indoor ventilation | [50, 51] |
| Contact rates and duration | [18, 52] |
| School and work calendars | [19, 20, 31] |

#### Appendix A.3. Relating environmental parameters to the seasonal *γ* **and collinearity between causal factors**

Differences in incoming solar radiation are a common cause of seasonal changes in temperate zones, having a major effect on all other environmental parameters, such as UV radiation, temperature and humidity, which show strong correlations with the cosine seasonality throughout the year.

Fig A.5 shows the correlations of the seasonal Γ parameter used in the transmission model, average insolation at the top of the atmosphere, temperature, and humidity, in four of the analysed countries. The data are from the NASA POWER project [53, 54]. Country areas were selected using a rectangular bounding box minimising border overlap.

High correlation coefficients such as 0.9 indicate that including these parameters as further explanatory variables in the current analysis would lead to strong multicollinearity. Consequently, when these parameters are used as independent explanatory variables in studies of seasonal effects, significant problems with multicollinearity are likely to create difficulties for causal inference. This poses a serious challenge for observational studies looking at individual seasonal and environmental factors and may be part of the explanation for previously inconclusive or contradictory results in the existing literature.

It is also worth noting that the different causal pathways described in Appendix A depend on environmental parameters with different timescales of aggregation. For example, indoor humidity in buildings is not an instantaneous function of outdoors humidity, temperature, and ventilation, but may exhibit some inertia if walls and plaster act as a reservoir of moisture, leading to averaging effects over timescales of days, weeks, and perhaps even months. While our holistic approach focuses only on the yearly oscillation, effectively integrating over all higher frequency causal effects, studies examining correlations between, e.g., daily average humidity and daily reproduction number are likely to miss some of the lower-frequency and delayed effects. When these longer timescale effects are included, the above described problem with multicollinearity becomes more pronounced.

**Figure A.5:**
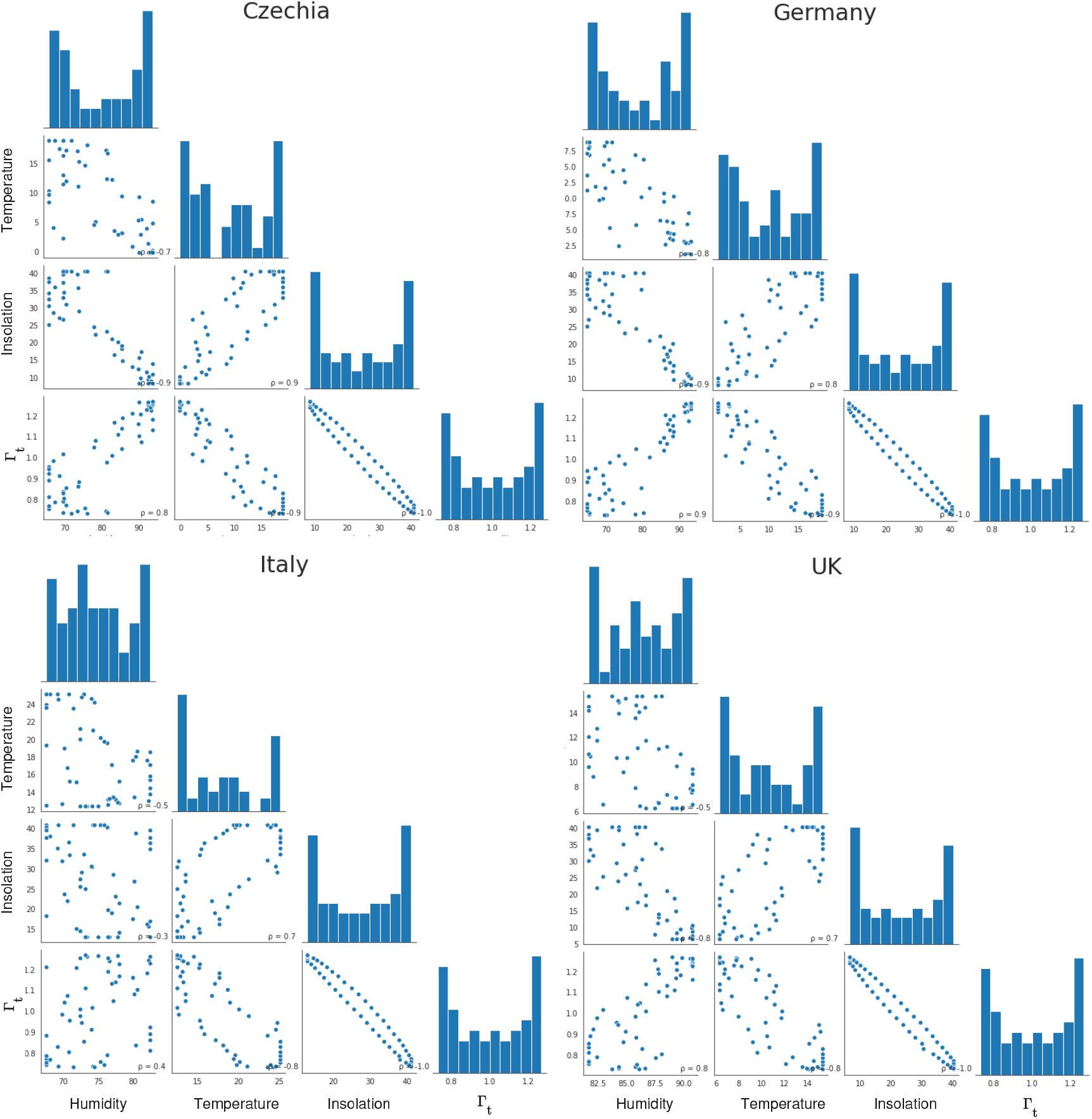
Correlations of the seasonal Γ parameter used in the transmission model, average insolation at the top of the atmosphere, temperature, and humidity. Each point is one weekly average value. *ρ* is the Pearson correlation coefficient.

### Appendix B. Datasets and implementation

Our Brauner et al. seasonal model implementation is based on the Brauner et al. code-base [23] and can be obtained at https://github.com/gavento/covid_seasonal_Brauner (version tag preprint-v2) together with the datasets used, including the temperate Europe dataset where we restrict the dataset of Brauner et al. [23] to the following 29 regions (out of 41 total):

*Albania, Andorra, Austria, Belgium, Bosnia and Herzegovina, Bulgaria, Croatia, Czech Republic, Denmark, Estonia, France, Germany, Greece, Hungary, Ireland, Italy, Latvia, Lithuania, Malta, Netherlands, Poland, Portugal, Romania, Serbia, Slovakia, Slovenia, Spain, Switzerland, United Kingdom*.

Our Sharma et al. seasonal model implementation is based on the Sharma et al. code-base [24] and can be obtained at https://github.com/gavento/covid_seasonal_Sharma/ (version tag preprint-v2). We use the Sharma et al. dataset without any modifications and with the same preprocessing, in particular, we also exclude data points with non-negligible prevalence of novel SARS-CoV-2 variants of concern.

### Appendix C. Detailed results

**Table C.2:** Inferred median values and confidence intervals of the seasonal amplitude *γ* and the peak-to-trough seasonality *R* reduction for temperate Europe countries.

| Model | Seasonal amplitude $\gamma$ | | |
| --- | --- | --- | --- |
|  | Median | 50% CI | 95% CI |
| Seasonal Sharma et al. | 0.275 | 0.248 – 0.300 | 0.200 – 0.348 |
| Seasonal Brauner et al. | 0.254 | 0.209 – 0.297 | 0.118 – 0.383 |
| Combined | 0.267 | 0.231 – 0.299 | 0.141 – 0.365 |

| Model | Peak-to-trough $R$ reduction [%] | | |
| --- | --- | --- | --- |
|  | Median | 50% CI | 95% CI |
| Seasonal Sharma et al. | 43.1 | 39.7 – 46.2 | 33.3 – 51.6 |
| Seasonal Brauner et al. | 40.5 | 34.6 – 45.8 | 21.1 – 55.4 |
| Combined | 42.1 | 37.5 – 46.1 | 24.7 – 53.4 |

**Figure C.6:**
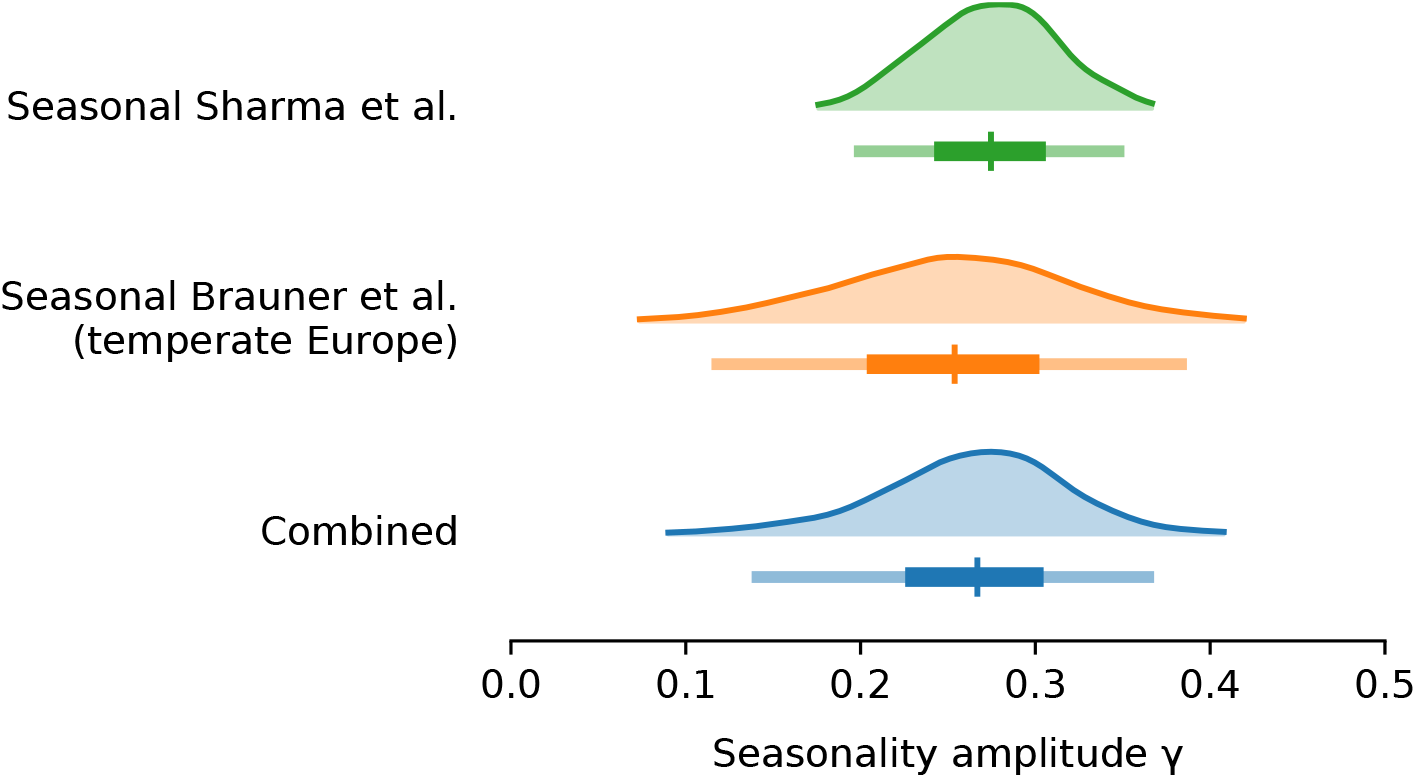
Posterior distributions the seasonal amplitude factor *γ* with 50% and 95% confidence intervals.

### Appendix D. Sensitivity to adjustment for mobility

Beyond NPIs and seasonality, voluntary changes in behaviour and contact patterns are important influences on the reproduction rate. As noted, seasonal variation in behavioural patterns such as time spent indoors is an important component of our holistic conception of seasonality (Appendix A). However, if there are behavioural changes over time that are causally unrelated with the transition between seasons, these may be mistakenly attributed to a causal effect of seasonal forcing.

**Figure D.7:**
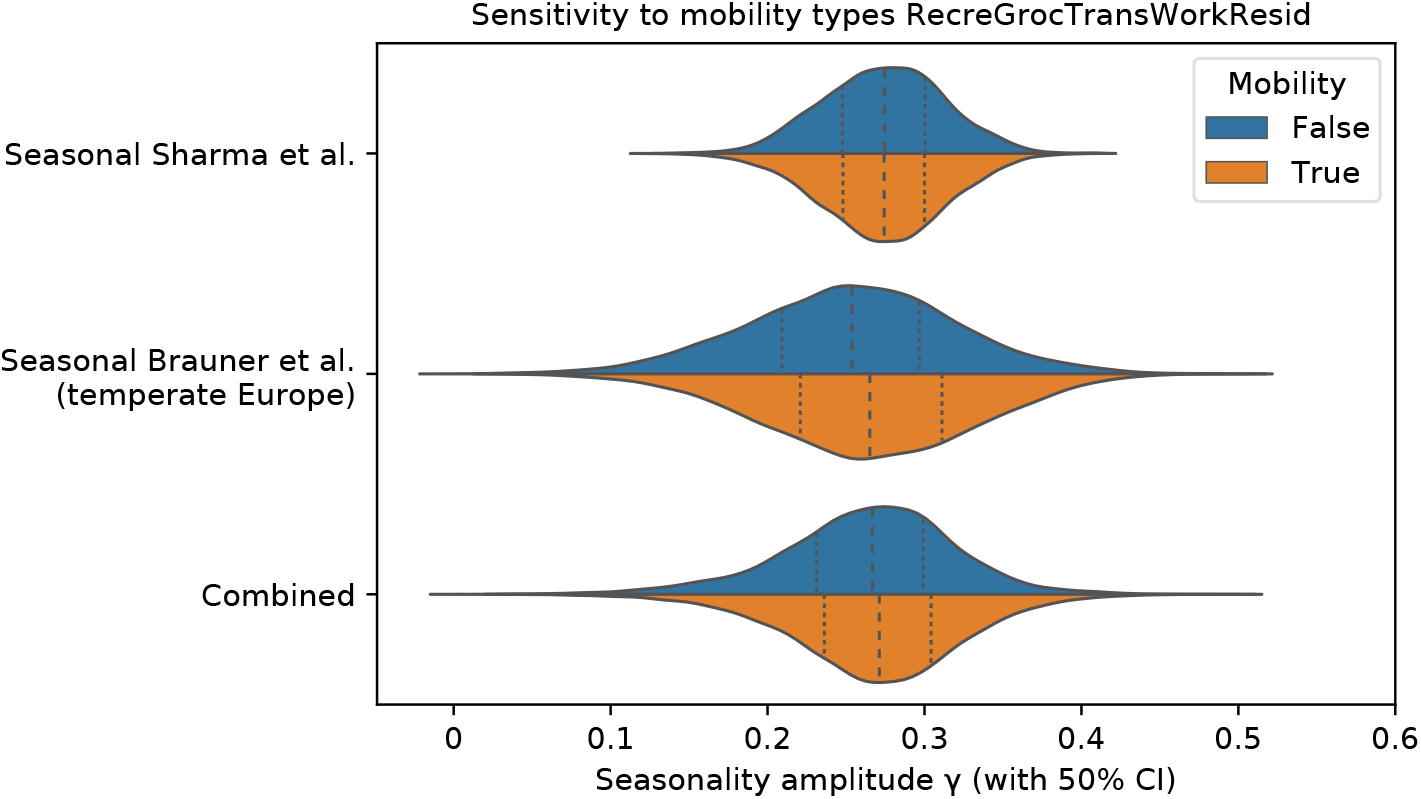
Comparison of posterior distributions of the seasonal amplitude *γ* in the main model vs with adjustment for mobility. Median and 50% confidence intervals.

To examine whether behavioural changes over the course of the pandemic biased our results, we incorporate mobility in the model by treating changes in mobility as a new distinct NPI “Mobility reduction” and run a full inference for NPIs and seasonality. We introduce the adjustment to mobility to both the Sharma et al. and the Brauner et el. models. For both models we use the country-level mobility data from Google Mobility Reports [55].

The Google Mobility Reports capture mobility change relative to a pre-pandemic, country-specific baseline in six categories: *Grocery & pharmacy, Parks, Transit stations, Retail & recreation, Residential, Workplaces*. The *Parks* category seems clearly causally related to seasonal and weather factors and we thus leave it out of the analysis.

Specifically, for the selected category set *C* above, each country *l*, and day *d*, we compute the activation of “Mobility Reduction” (MR) NPI as

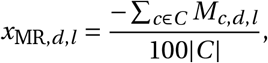

linearly mapping the pre-pandemic mobility level in each country to 0.0 and the (hypothetical) zero mobility to 1.0.

We find that our seasonality estimates are robust to adjusting for a combined mobility trend in locations. Specifically, our combined median estimate of *γ* is 0.2712 with mobility vs 0.2668 with the default model, a difference of 1.64%. See Figure D.7 for posterior distributions of *γ*.

### Appendix E. Random walk noise comparison

The Sharma et al. model contains a random walk process on log *N*_*t,l*_, the logarithm of a multiplicative factor in *R*_*t,l*_, in order to account for continuous slow changes of *R*_*t*_ through unobserved external factors such as unobserved NPIs or environmental transmission factors [24].

**Figure E.8:**
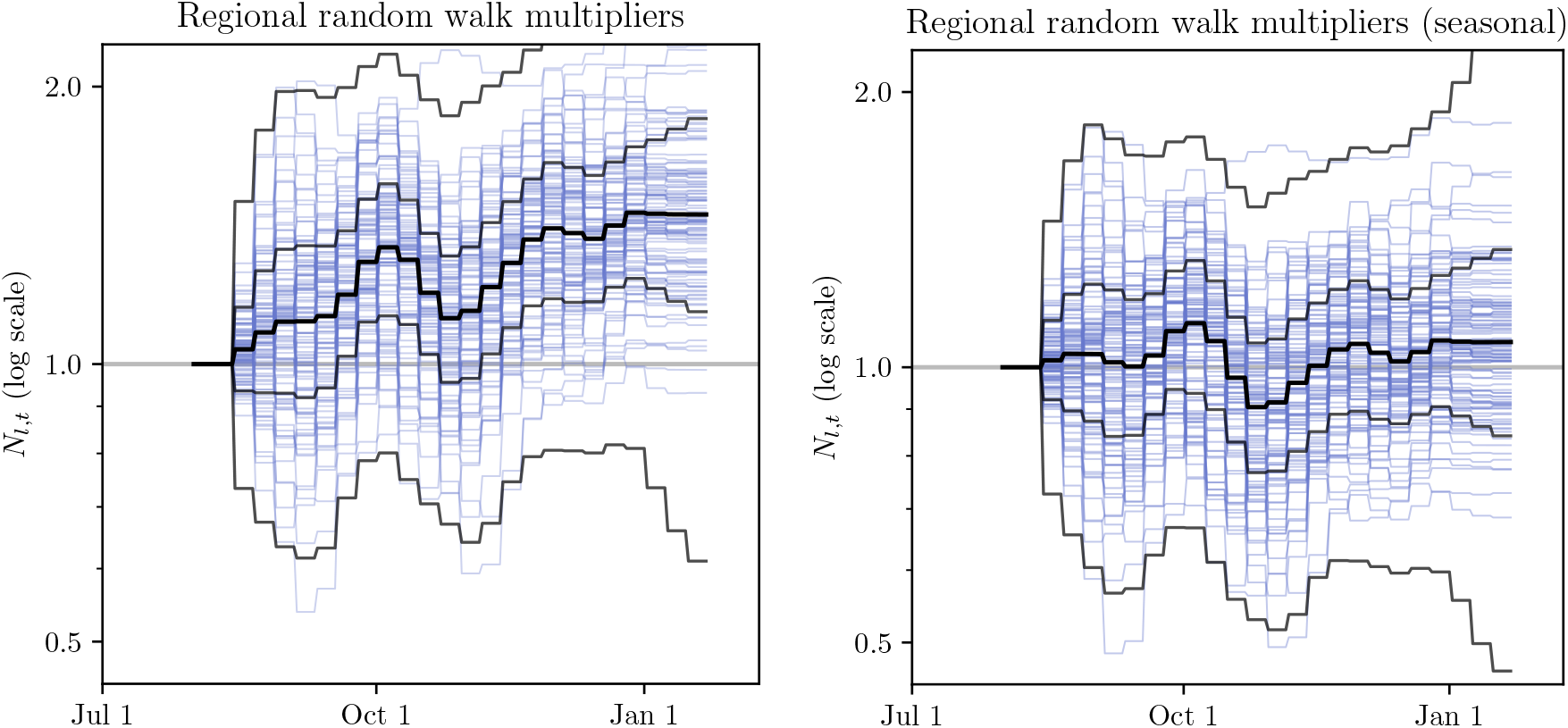
Inferred *N*_*t, l*_ for non-seasonal (left) and seasonal (right) models. Blue lines are median *N*_*t, l*_ for each region, black bands indicate median, 50% CI and 95% CI.

To show the effect of modelling seasonality on the inferred random walk noise, we compare *N*_*t,l*_ for the two models in Figure E.8. While the random walk trajectories are comparable in width, the median trajectory of the non-seasonal model follows an increasing trend. Note that the random walk multiplier is modelled as a symmetrical random walk in log-space. Also note that the Sharma et al. model allows only weekly changes in *N*_*t,l*_.

To quantify the improvement, we compute the mean squared deviation (MSD) of log *N*_*l,t*_, i.e. the random-walk multiplier in log-space, across the sampled random walks. We find this MSD to be 0.131 for the non-seasonal model, and 0.072 for the seasonal model, a 45% decrease. Note that here we compute MSD as

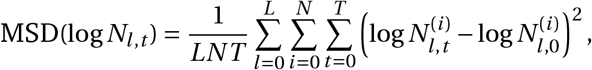

where 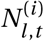 is the *i* -th sample of *N*_*l,t*_.

**Figure E.9:**
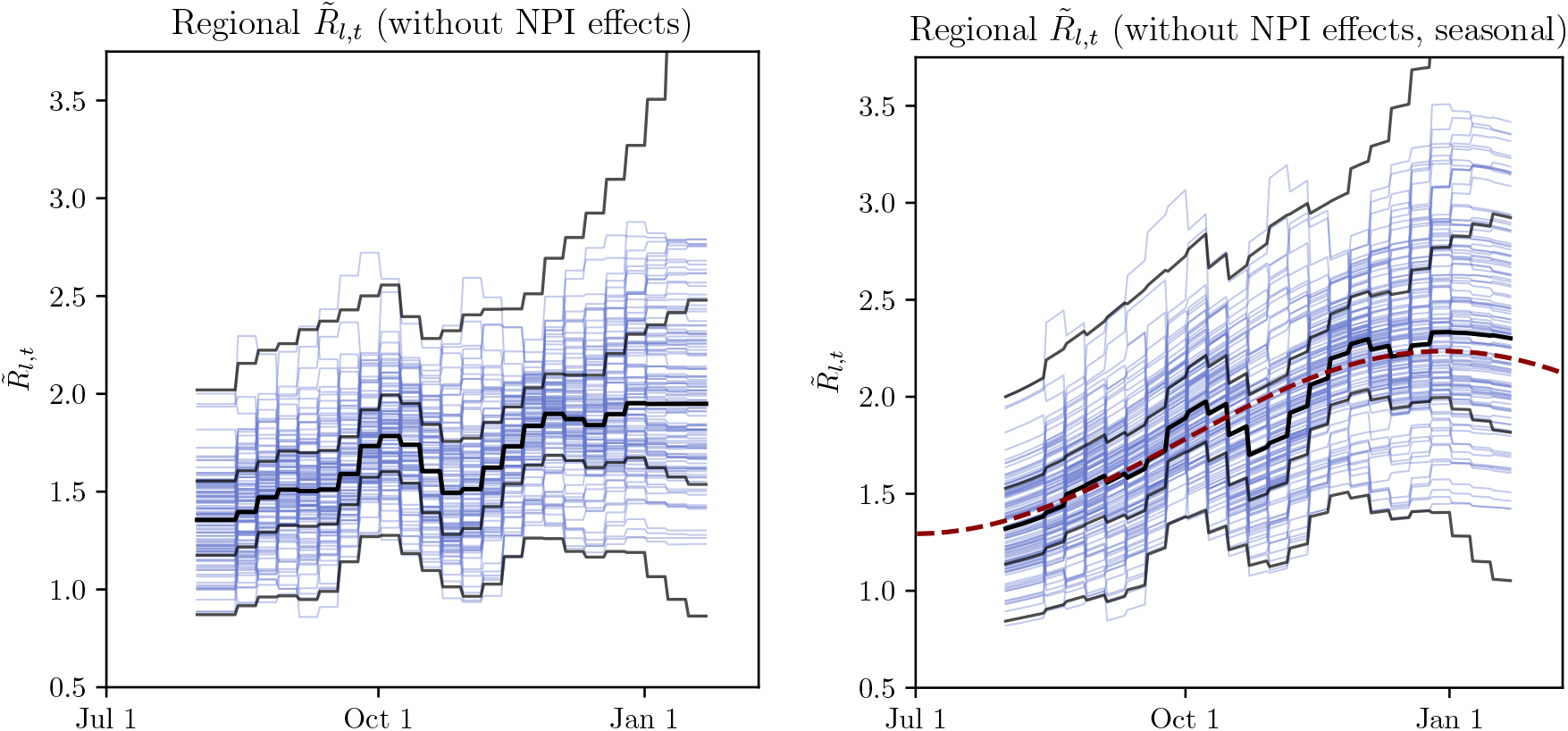
Inferred 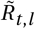 for non-seasonal *(left)* and seasonal *(right)* models. Blue lines are median 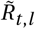 for each region, black bands indicate median, 50% CI and 95% CI. Red dashed line shows cosine seasonality with the inferred amplitude *γ ≈* 0.267 applied to median inferred *R*_0_.

We compare 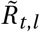 for the two models – the reproduction factor derived from region-specific *R*_0_ by the random walk process and by seasonality effect (in the seasonal model) but *before* applying transmission reduction of the active NPIs:

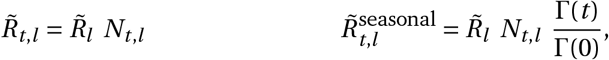

where *N*_*t,l*_ is the random walk noise.

Figure E.9 illustrates how the inferred 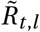 are comparable for the non-seasonal and seasonal model. However, the non-seasonal model random walk is of a larger overall amplitude and has an asymmetric trend compared to the seasonal model, as shown on Figure E.8. We interpret this as an indirect evidence towards seasonality improving the quality of model fit on Sharma et al. data.

Note that the noise terms used in Brauner et al. are of different type and the model does not contain a comparable random walk noise term.

### Appendix F. Sensitivity analysis

#### Appendix F.1. Inferring the peak seasonality day

To examine the fitness of our seasonality peak estimation, we place a prior of *𝒩* (Jan 1, 45^2^) on *d*_*γ*_ instead of a fixed date. Figure F.10 shows the distribution of the seasonality multiplier cosine curves Γ(*t*) inferred with prior on *d*_*γ*_. Figure F.11 shows both the inferred seasonality peak day *d*_*γ*_ and the seasonality amplitude *γ*.

Note that the estimated *d*_*γ*_ are shown as a model validation, illustrating the range of seasonality peak the models and data are consistent with – we do not claim the models and data can infer the peak with accuracy. Note that the inferred *γ* in the model inferring the seasonality peak is virtually unchanged relative to a fixed seasonal peak day model (Figure F.11 bottom).

**Figure F.10:**
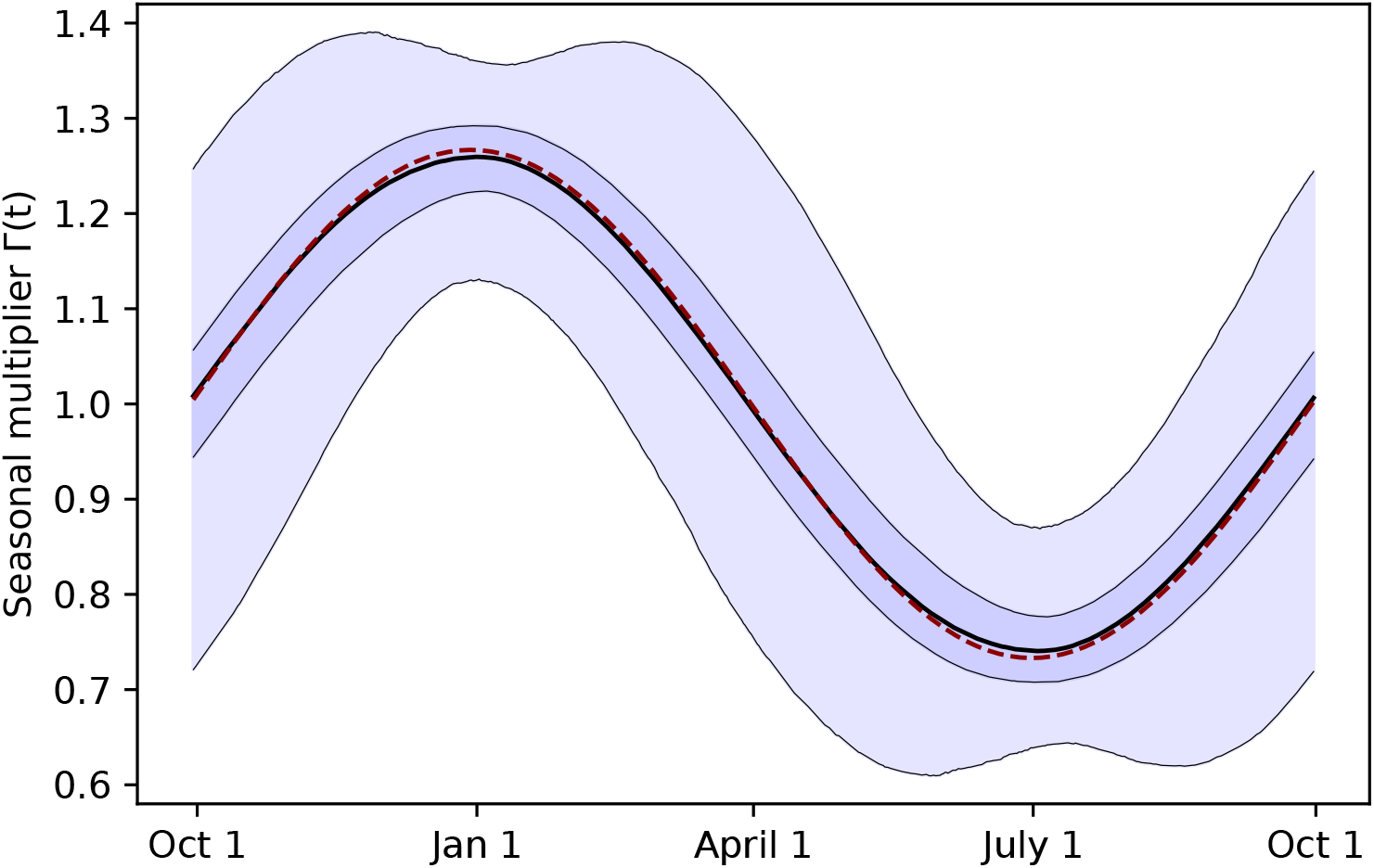
Distribution of Γ(*t*) for the combination of Sharma et al. and Brauner et al. seasonal models with a prior on *d*_*γ*_, with median and 50% and 95% CIs. The underlying Γ(*t*) curves are parameterized by the joint posterior distributions on *γ* and *d*_*γ*_. The dashed red line is the median Γ(*t*) inferred with fixed *d*_*γ*_ *=* Jan 1 for comparison.

**Figure F.11:**
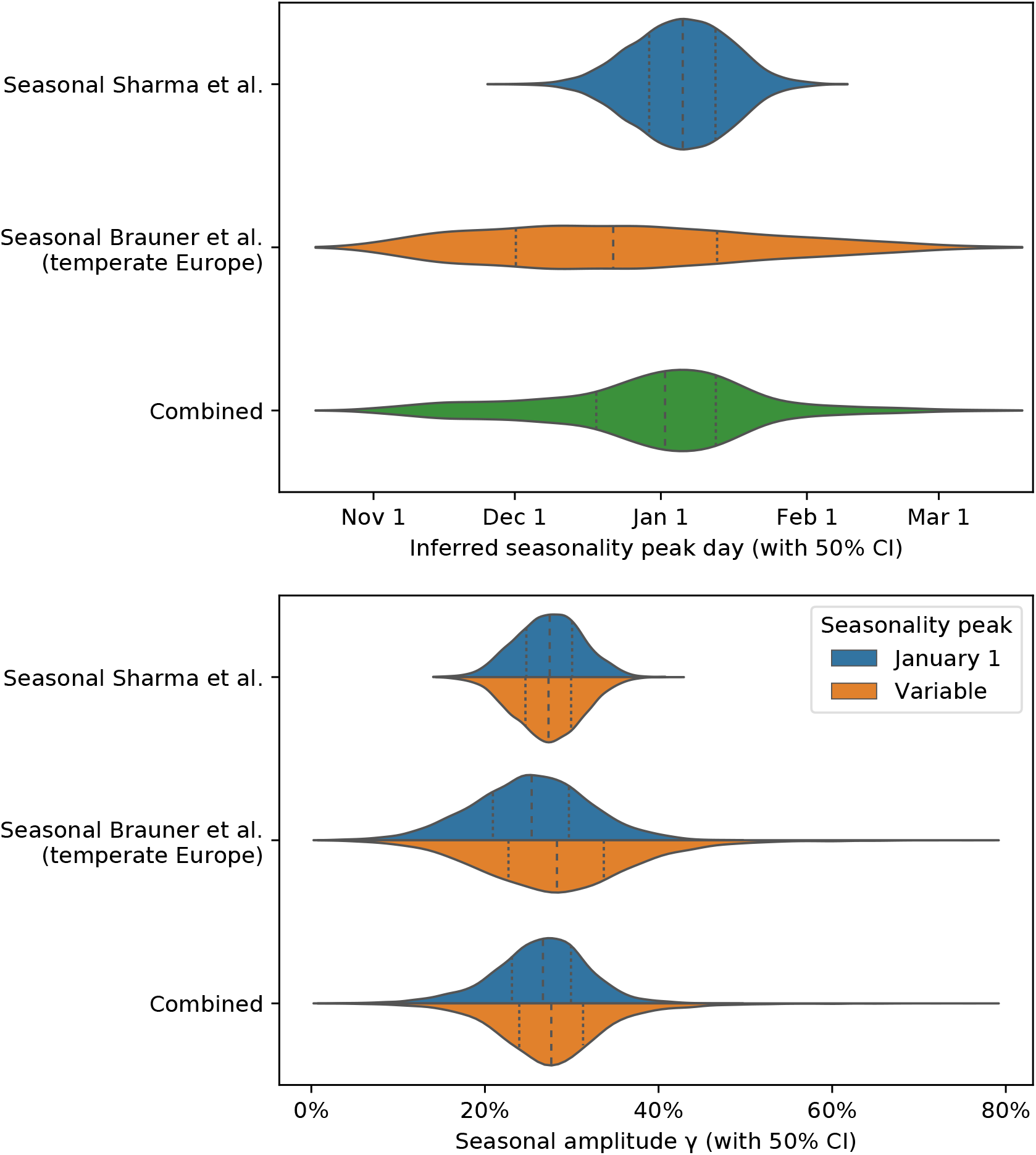
Inferred peak seasonality day *d*_*γ*_ *(top)* and *γ* posterior comparison in models with the peak day fixed *vs* with a variable peak day with *𝒩* (Jan 1, 45^2^) prior (bottom).

#### Appendix F.2. Sensitivity to peak seasonality day

We test model sensitivity to the choice of peak seasonality day *d*_*γ*_ for *d*_*γ*_ ∈ {Dec 4, Dec 18, Jan 1, Jan 15, Jan 29, Feb 12, Feb 26}. We observe that the inferred combined effect of the NPIs and the inferred seasonality are stable for *d*_*γ*_ in December and January with the exception of the combined NPI effect in Brauner et al. Note that Sharma et al. is particularly robust in this range.

**Figure F.12:**
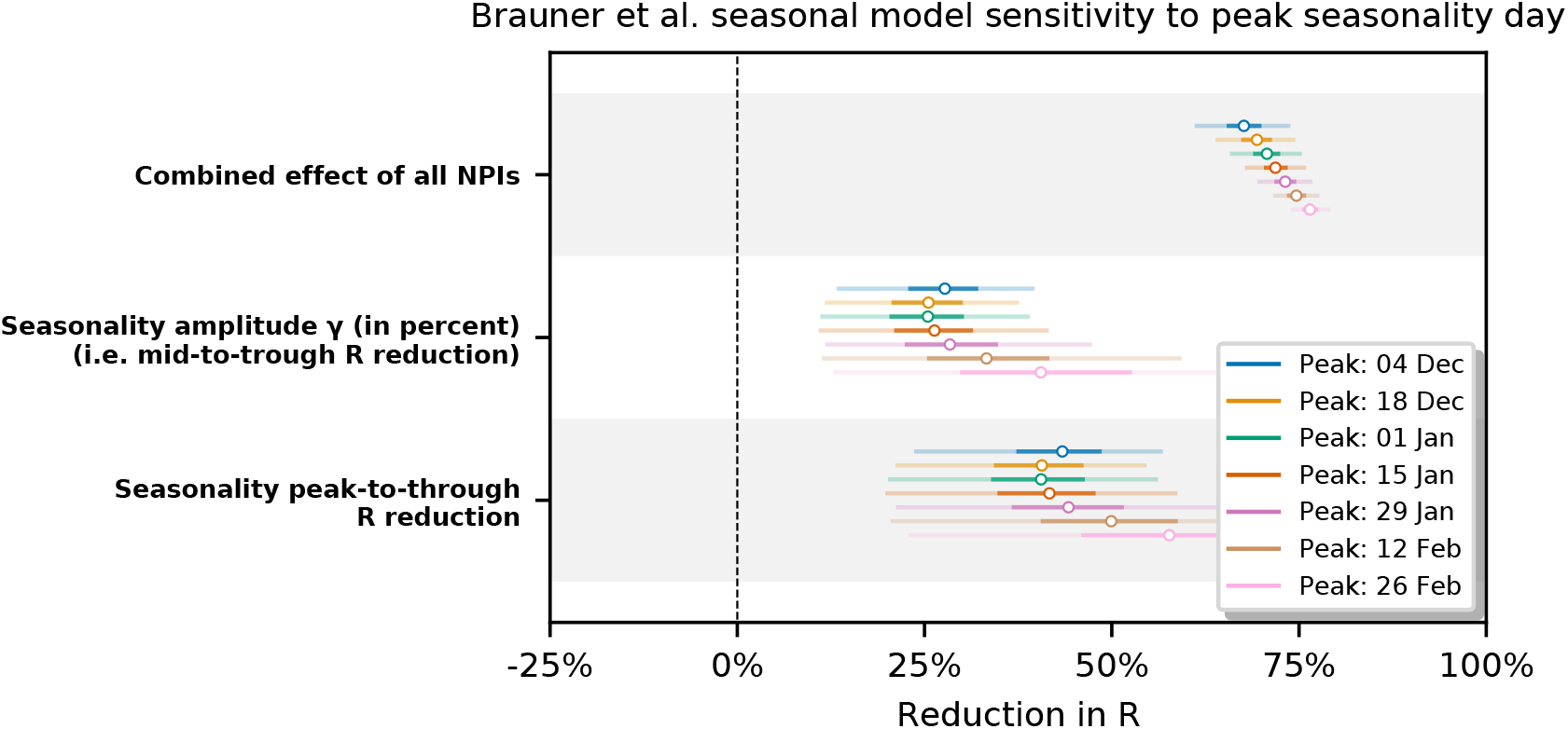
Sensitivity of Brauner et al. seasonal model to the choice of *d*_*γ*_.

**Figure F.13:**
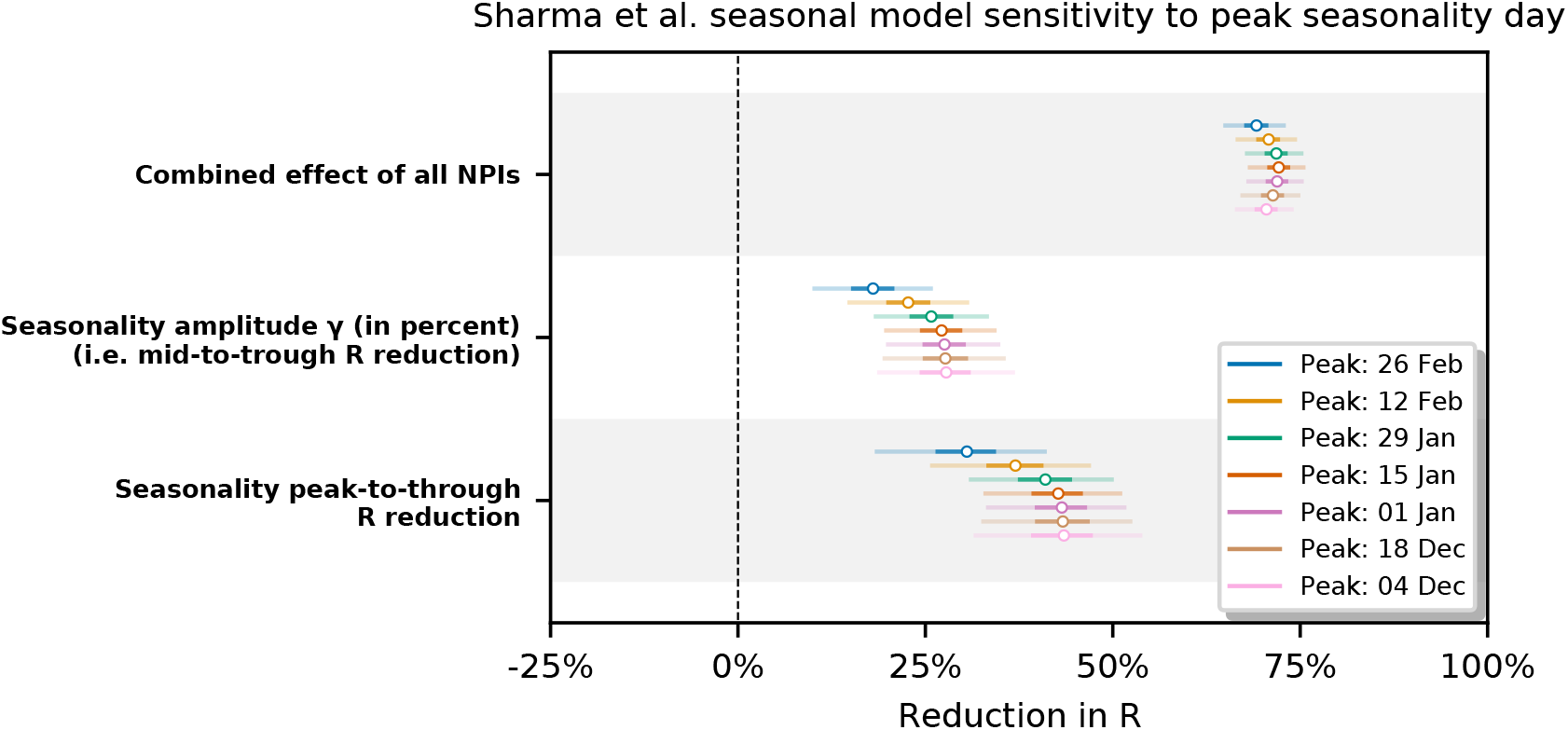
Sensitivity of Sharma et al. seasonal model to the choice of *d*_*γ*_.

#### Appendix F.3. Sensitivity to initial *R*_0_ **prior**

We test our model sensitivity to the choice of the mean of the initial 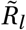 prior (i.e. location-specific *R*_0_ on the first day of the dataset). We analyse the 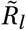 prior mean in ranges similar to the sensitivity analyses in Sharma et al. [24] and Brauner et al. [23].

**Figure F.14:**
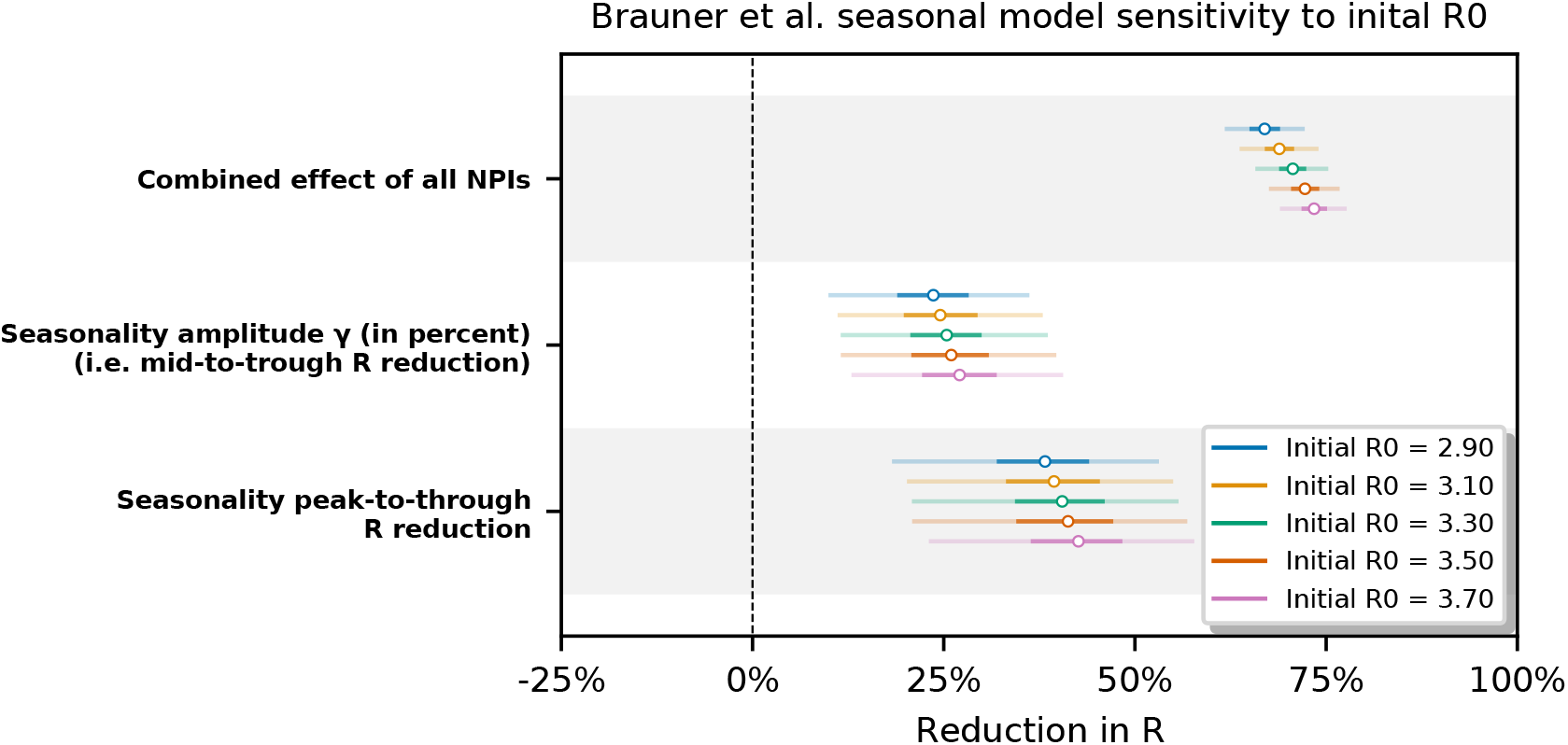
Sensitivity of Brauner et al. seasonal model to the initial *R*_0_ prior mean.

**Figure F.15:**
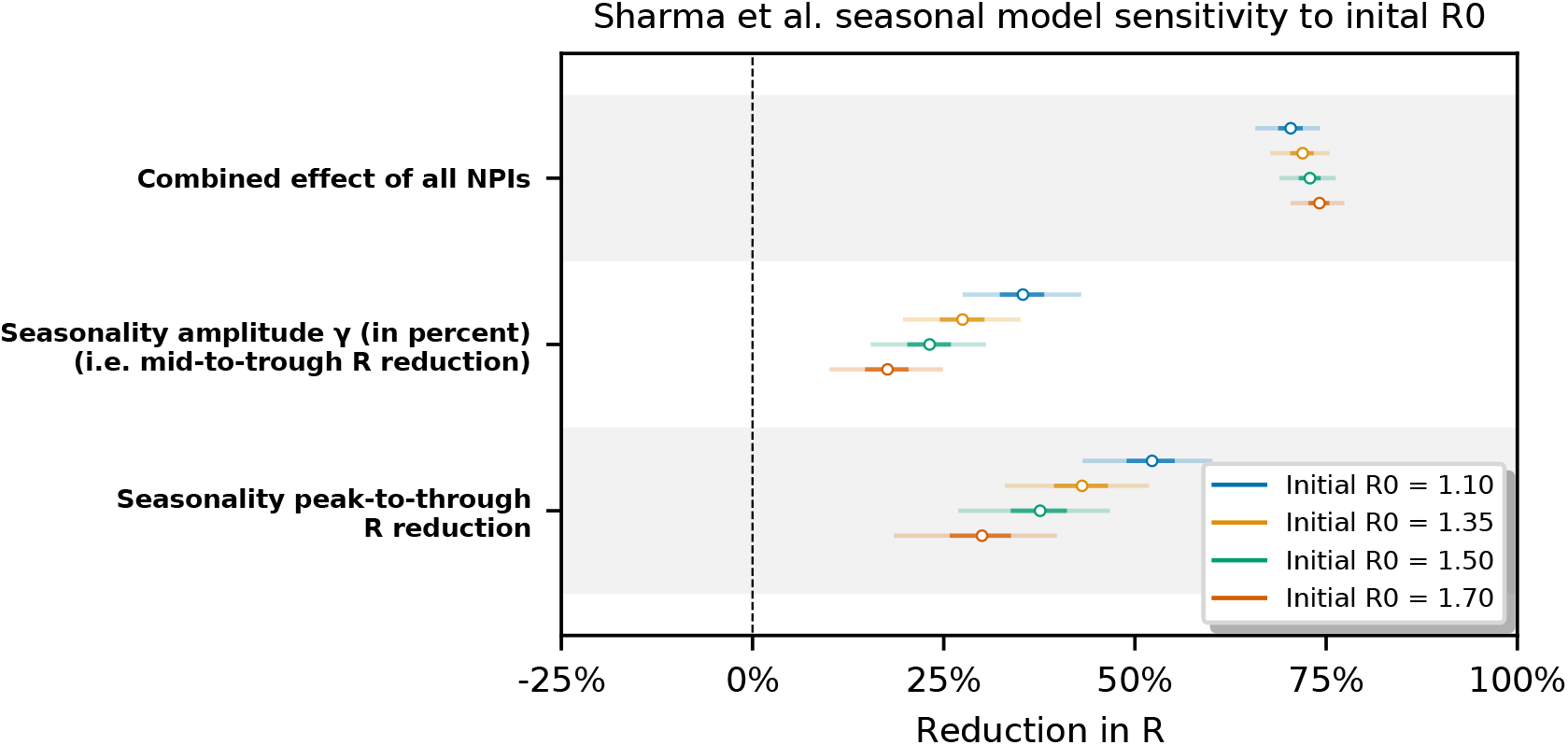
Sensitivity of Sharma et al. seasonal model to the initial *R*_0_ prior mean.

This analysis is motivated by the seasonal amplitude parameter *γ* being closely connected with 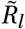 via Equations (1) and (3). Mis-specifying the initial 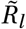 could be compensated for by the model e.g. by a different amplitude *γ* and therefore also the slope of Γ(*t*) in the seasonality multiplier sine curve.

In Figures F.14 and F.15 we observe the inferred combined effect of the NPIs and the inferred seasonality to be mostly stable in the Brauner et al model. However, in the Sharma et al. seasonal model the inferred seasonality amplitude and peak-to-trough reduction are mildly sensitive to the 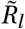 prior. (Note that both those parameters are very closely tied together.)

Note that both original models also exhibit some sensitivity of the effect of NPIs to *R*_0_ prior mean; see Figure S11 in Supplementary material of Brauner et al. [23] and Figure S13 in Supplement of Sharma et al. [24] (v1).

#### Appendix F.4. Inferred total NPI effects in various models

We compare the inferred total NPI effect in different models and data subsets to verify its stability: seasonal vs non-seasonal (original) models, and the original full dataset vs the dataset restricted to temperate Europe countries (“TE” in plot for Brauner et al. model).

We observe that restricting the dataset regions in Brauner et al. model has very little effect on the inferred combined NPI effect.

Switching to a seasonal model produces a small decrease (resp. increase) in combined NPI effect in Brauner et al. (resp. Sharma et al.) model. While this may be coincidental, this effect is consistent with a hypothesis that a part of the seasonality-related change in *R*_0_ (i.e. the proposed spring decrease of *R*_0_ in Brauner et al., fall increase in Sharma et al.) is in part attributed to NPI activations in both models. Recall that Brauner et al. only considers NPI activations in their model, and Sharma et al. dataset is dominated by NPI activations compared to deactivations. However note that both models do contain noise terms for growth rate and other mechanisms to model small or slow changes in *R* due to unobserved factors, so the extent of this effect remains unclear.

**Figure F.16:**
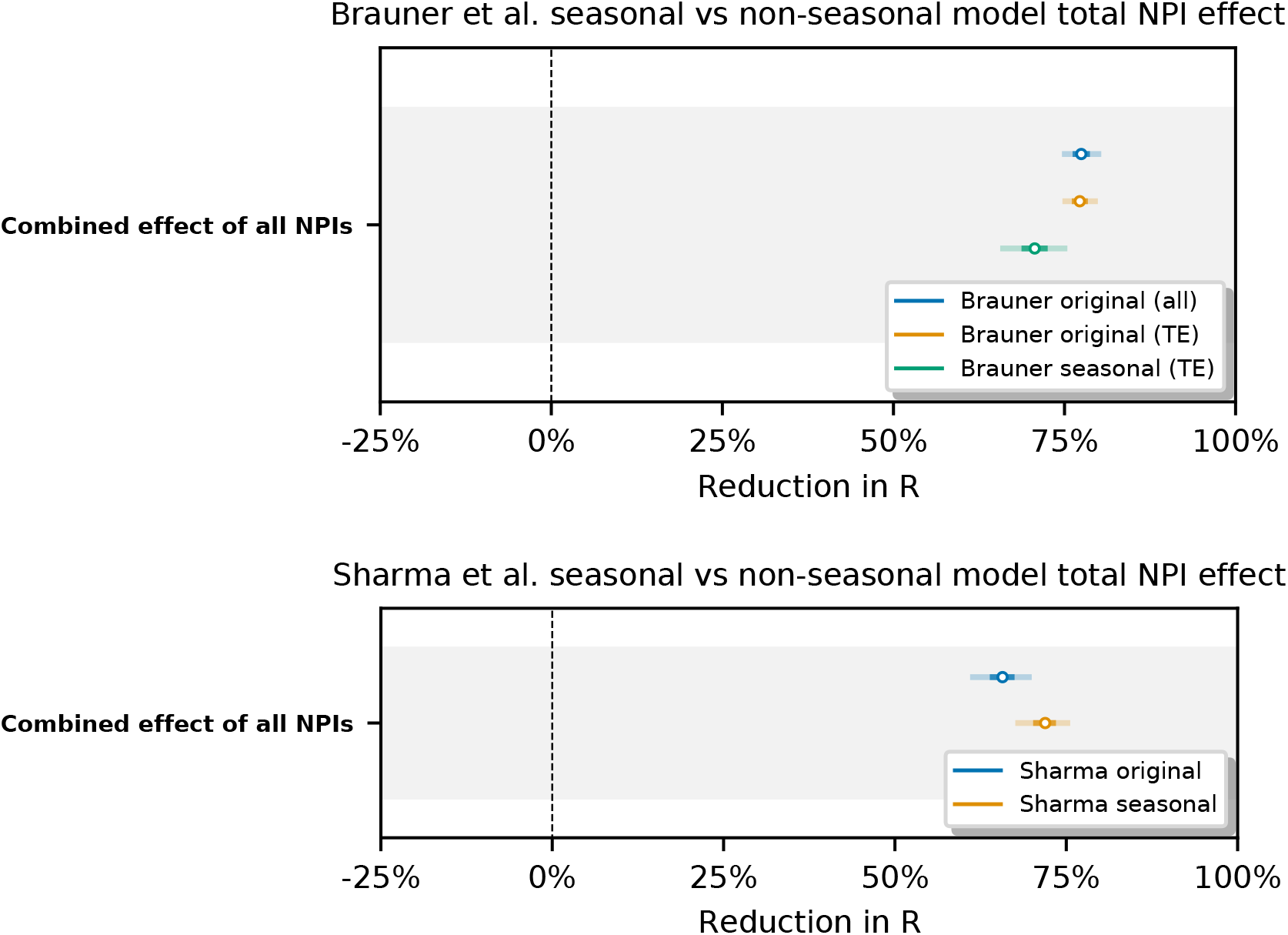
Inferred NPI effects on Brauner et al. dataset across several models. Runs marked with “TE” are restricted to temperate Europe countries, runs marked with “all” are ran on the original Brauner et al. dataset.

## Footnotes

1 Sinusoidal seasonality is well-defined only for amplitudes −1 ≤ *γ* ≤ 1. We restrict it to −0.95 ≤ *γ* ≤ 0.95 to improve model stability.

